# Nowcasting and Forecasting the Spread of COVID-19 and Healthcare Demand in Turkey, A Modelling Study

**DOI:** 10.1101/2020.04.13.20063305

**Authors:** Şeyma Arslan, Muhammed Yusuf Özdemir, Abdullah Uçar

**Author notes:** Contributed equally.

## Abstract

**Background:** This study aims to estimate the total number of infected people, evaluate the effects of NPIs on the healthcare system, and predict the expected number of cases, deaths, hospitalizations due to COVID-19 in Turkey.

**Methods:** This study was carried out according to three dimensions. In the first, the actual number of infected people was estimated. In the second, the expected total numbers of infected people, deaths, hospitalizations have been predicted in the case of no intervention. In the third, the distribution of the expected number of infected people and deaths, and ICU and non-ICU bed needs over time has been predicted via a SEIR-based simulator (TURKSAS) in four scenarios.

**Results:** According to the number of deaths, the estimated number of infected people in Turkey on March 21 was 123,030. In the case of no intervention the expected number of infected people is 72,091,595 and deaths is 445,956, the attack rate is 88 · 1%, and the mortality ratio is 0·54%. The ICU bed capacity in Turkey is expected to be exceeded by 4·4-fold and non-ICU bed capacity by 3·21-fold. In the second and third scenarios compliance with NPIs makes a difference of 94,303 expected deaths. In both scenarios, the predicted peak value of occupied ICU and non-ICU beds remains below Turkey’s capacity.

**Discussion:** Predictions show that around 16 million people can be prevented from being infected and 94,000 deaths can be prevented by full compliance with the measures taken. Modelling epidemics and establishing decision support systems is an important requirement.

## Introduction

Infectious diseases can persist in certain populations (endemic), spread at a sudden rate and affect wider populations (epidemic), or turn into a global threat (pandemic) as in the 1918 Spanish flu.^1^ Coronaviruses, which were first detected in 1960, have been observed in humans to date and have seven subtypes, and are responsible for the SARS outbreaks in 2003 and MERS in 2012.^2^

A new type of coronavirus (later named Sars-Cov-2) first drew attention on 31 December 2019 after 27 pneumonia cases with unknown etiology were detected in Wuhan, China and reported to the World Health Organization (WHO).^3,4^ The epidemic caused by the virus, called COVID-19, spread rapidly between countries and continents and was ultimately considered pandemic by the WHO on 11 March 2020.^5^

The rapid progression of the COVID-19 pandemic and its devastating effects in many countries has revealed the vital nature of epidemic modelling studies to evaluate the course of the epidemic and its burden on countries’ health systems properly. Stochastic, deterministic and agent-based models have been used in the scientific literature to model the spread of COVID-19.^6,7^

Turkey has also taken precautions due to the COVID-19 pandemic and many additional measures were implemented after the identification of the first national case on 11 March 2020.^8^ These measures include Non-Pharmaceutical Interventions (NPIs) such as school closures, cancellation of arts and sports events, mandatory quarantine for people who have travelled to the country from abroad, the general closure of public places such as cafes/cinemas/wedding halls, making mask usage in grocery stores obligatory, curfew for the citizens over the age of 65 and under 20 and those with chronic illnesses.^9-11^

This study aims to estimate the total number of infected people, evaluate the consequences of social interventions on the Turkish healthcare system, and predict the expected number of cases, intensive care needs, hospitalizations and mortality rates in Turkey according to possible scenarios via a modified version of the SEIR-based outbreak modelling method. Thus, it aims to contribute to the pandemic response policies adopted in Turkey by providing an epidemiological framework.

## Materials and Methods

### 1. Study Design

This study was carried out according to three different dimensions. In the first dimension, the actual number of people infected in the community was estimated using the number of deaths in Turkey. In the second dimension, the expected total numbers of infected people, deaths, hospitalizations, and intensive care unit (ICU) bed needs were predicted in the case of no intervention.

The predictions in the second dimension include cumulative numbers only. Thus, additional calculations were required to predict the distribution of healthcare needs, patients, and deaths over time. Therefore, a third dimension was added to the study to model the distribution of the expected numbers to determine the health resources required based on this model, and to predict the impact of social interventions on the progression of the epidemic.

In this third dimension, the SEIR model was used for estimations and predictions. This model divides society into four main compartments during the epidemic: those who are not yet infected (Susceptible), those who have been exposed to the agent but show no signs of infection (Exposed), those who have had symptoms of the disease (Infectious), those whose who have either recovered or died from the disease (Removed).^12^

### 2. First Dimension Assumptions and Forecasting Algorithm

The ratio of deaths in the total infected population is identified in the literature as the Infection Fatality Ratio (IFR).^13^ There may be a time shift bias in the estimations based on the number of deaths. For more accurate estimates, the number of deaths observed on a given day should not be compared to the number of infectious people for the same day; instead, it should be compared to the day the infection started.^14^ Thus, in this dimension of the study, the number of infected people was estimated by using the number of deaths based on IFR. According to the studies, the time that elapses from initial symptoms to death is about 18 days.^13^ The number of infected people was estimated according to a delay of 18 days, and the remaining days were projected with a quadratic growth curve which has the highest R^2^ value (0·9936). This study used the average IFR (0·66% [0·39-1·33]) and age-specific IFR values which were adjusted for the United Kingdom and the United States in Imperial College London (ICL) modelling based on calculations by Verity et al.^13^

### 3. Second Dimension Assumptions and Forecasting Algorithm

The COVID-19 overall infection rate for Turkey was considered to be 81%.^15^2018 TurkStat census data was used for age stratification. Using the expected age-specific hospitalization and intensive care ratios, total hospitalization numbers and ICU needs are estimated for each age group. First dimension values were used for IFR values. By applying age-specific IFR values to the expected number of infected people in the relevant age group, the highest number of expected deaths was determined.^13,15^ In this dimension, it was assumed that no measures were taken, and the pandemic is free to spread throughout society.

### 4. Third Dimension Assumptions and Forecasting Algorithm

In this dimension of the study, a SEIR-based model was created, and a simulator called TURKSAS was developed by adding transmission dynamics as well as clinical dynamics and NPIs dynamics. The TURKSAS model structure is as presented in Figure 1.

**Figure 1:**
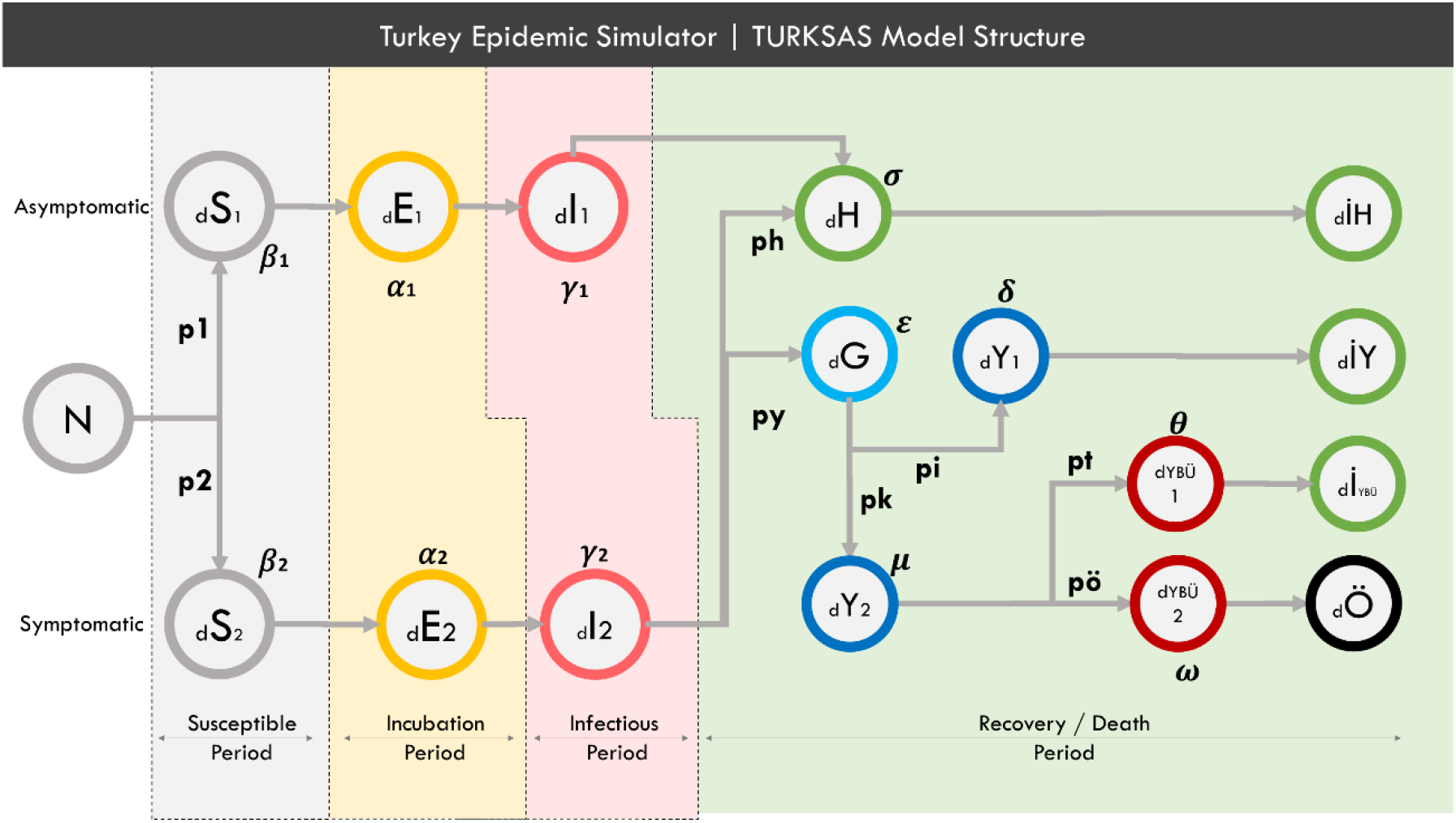
TURKSAS model structure. In a time section; N: Total population; d: delta (expressing the change of the related cluster over time) S- E- I: The number of Susceptible-Exposed-Infected people, respectively, in the relevant time section. H: Infected who have mild symptoms. İH: Those who have recovered with mild symptoms. G: Infected and have not yet applied to the hospital. Y: Infected who apply to the hospital and have occupied non-ICU beds. IY: Those who have recovered in hospital and been discharged. Iybu: Those who have recovered from ICU. YBÜ1: Those who will recover in ICU. YBÜ2: Those who will die in ICU. Ö: Those who have died. For other parameters, see Mathematical Equation of the Model.

#### • Mathematical Equation of the Model

**1*st Group*:** *Asymptomatic cases*

**2*nd Group*:** *Symptomatic cases*

***(p*:** *proportion* | ***ICU*:** *Intensive Care Unit)*

***p*_1_:** *Asymptomatic case proportion*

***p*_2_:** *Symptomatic case proportion*

***py*:** *Symptomatic and will apply to the hospital*

***ph*:** *Symptomatic and will have mild disease*

***pi*:** *will recover from the hospital*

***pk*:** *will need ICU Bed*

***pt*:** *will recover from ICU*

***p*ö:** *Fatality rate among ICUs according to IFR*

***R*0:** *Number of people contaminated by an infected*

***T_inc_*:** *Incubation pediod*

***T_inf_*:** *Infectious period*

***S*:** *Susceptible*

***E*:** *Exposed*

***I*:** *Infectious*

***H*:** *Mild cases*

**İ*H*:** *Recovered with mild symptoms*.

***G*:** *Infected but have not yet applied to the hospital*

***Y*_1_:** *Applied to the hospital and will recover*

***Y*_2_:** *Applied to the hospital and need ICU*

**İ*Y*:** *Recovered from the hospital without ICU need*

Ö: *Died*

***YB***Ü**1:** *Still in ICU and will be recovered*

İ***yb***ü: *Recovered from ICU*

***YB***Ü**2:** *Still in ICU and will die*

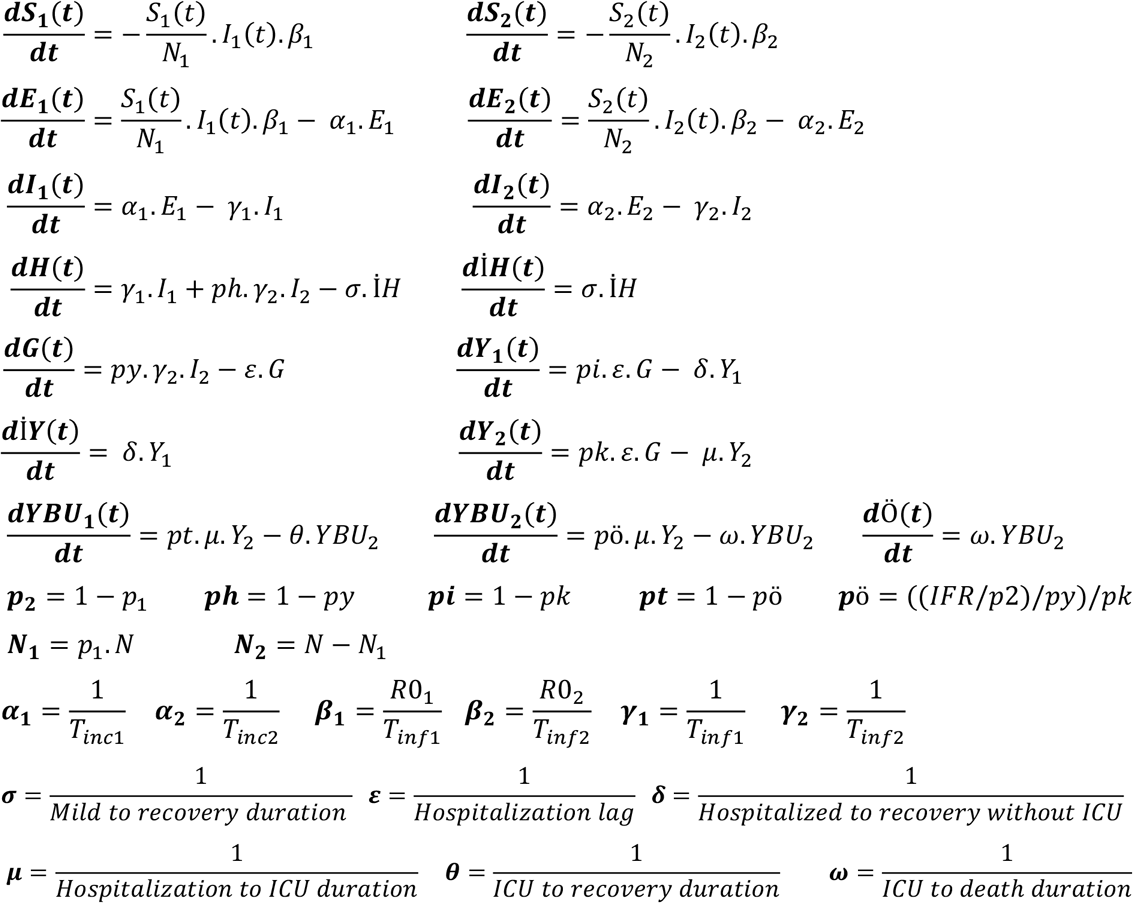

Because the incubation period, infectious period and R0 variables differ between symptomatic and asymptomatic cases, these two groups are considered to be separate community layers within this model. Also, it is assumed that asymptomatic cases will not apply to the hospital and die. The R compartment was also restructured to predict the need for health care. Some infected people will recover with mild symptoms without requiring hospital admission (H). Some will be late to apply to the hospital even though they show symptoms (G). After the delay, these people will apply to the hospital (Y). It is assumed that all positive cases admitted to the hospital will initially be transferred to the non-ICU beds. Some of these patients will recover directly from the service (İY) and some will recover and be discharged from ICU (YBU1). Others will go to ICU (YBU2) and then die (Ö).

Due to a lack of studies that estimate the local clinical care dynamics and durations in Turkey, we used coefficients and assumptions from various scientific studies.

#### • Transmission Dynamics

The average incubation period was accepted to be 4·6 days for asymptomatic cases and 5·1 days for symptomatic cases, and the infectiousness period was accepted to be 6·5 days for both groups.^15,16^ Symptomatic cases were considered to be two times more infectious than asymptomatic.^15^ It is assumed that R0 values are between 2 and 3 for Turkey.^17,18^ Considering that the study on the *Diamond Princess* (cruise ship) was close to a prospective cohort design, the rate of asymptomatic cases was accepted to be 17·8% in our study.^19^

#### • Clinical Dynamics

It has been assumed that people with mild symptoms will not apply to the hospital and their recovery will take 22 days.^20^ The delay time for hospital admissions is considered to be 5 days, and the period from hospitalization to recovery is considered to be 10 days.^21^ The duration of recovery from ICU to discharge is considered to be 15 days, and the duration from ICU to death is considered to be 7 days.^22,23^ Duration for referral to ICU after hospitalization was assumed to be 5 days from expert opinion. The duration from the onset of symptoms of the disease to death is considered to be 17·8 days.^13^ The total ICU bed and non-ICU bed capacity of Turkey is considered to be 38,098 and 193,095, respectively.^24^

#### • NPIs Dynamics

NPIs decrease the number of contacts, which accordingly decreases the value of R0 directly. This decrease affects all outputs over the β value in the equation. The impact of social interventions on the R0 value in European countries is presented in detail in the ICL March 30 report.^25^ In TURKSAS, the impact values from the ICL report were used and simulations were made specific to the dates when each intervention was activated. It was also calculated the extent to which the social interventions applied in Turkey reduced the default R0 value in the model over time. The dates the NPIs were applied, relative percentage reduction in R0, and assumptions about social compliance to NPIs in Turkey are presented in Table 1.

**Table 1:**
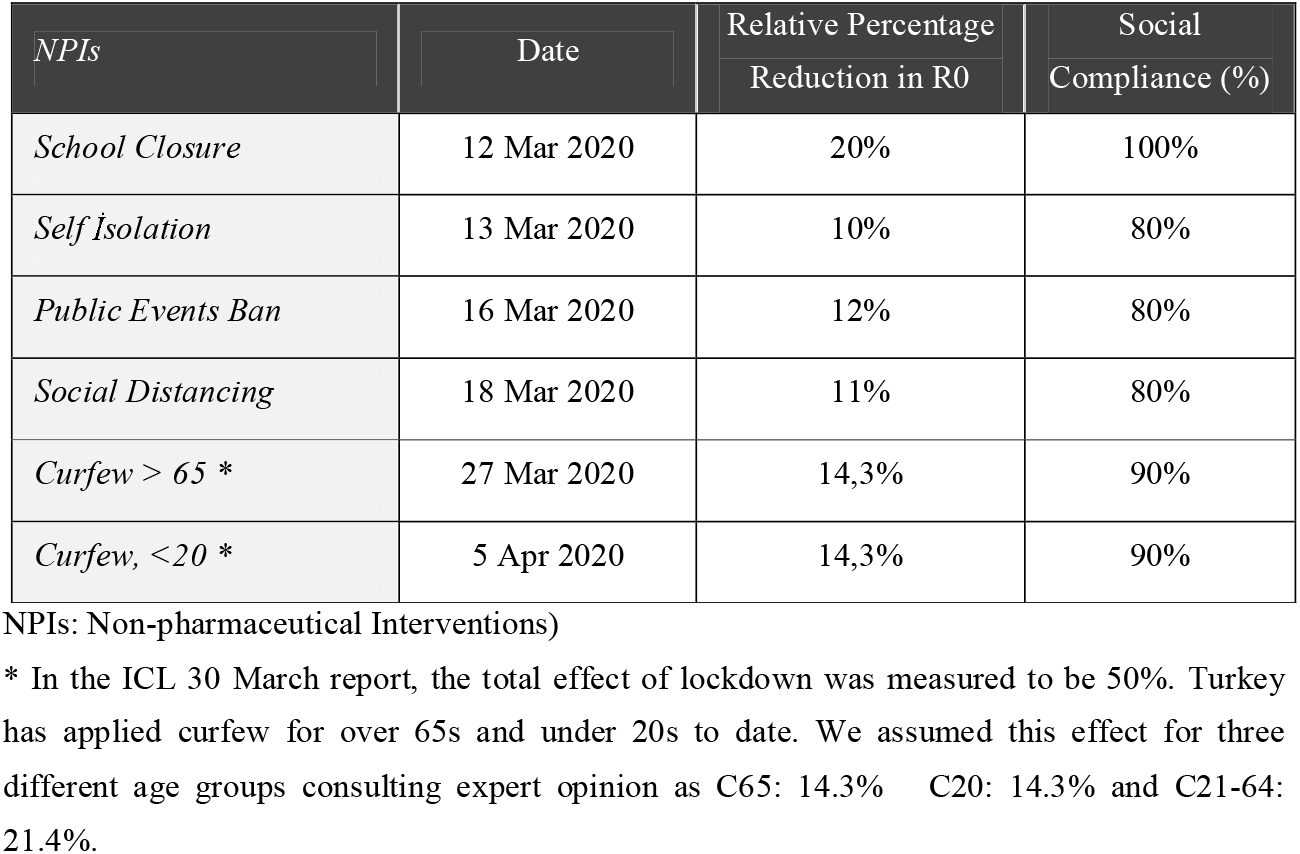
Effect of NPIs on R0 value (8) and assumptions regarding social compliance with policies (NPIs: Non-pharmaceutical Interventions)

## Results

### 1. First Dimension

According to the estimates based on the number of deaths (announced daily), the number of infected people on March 17 was 75,909. The number of infected people in society according to IFR and the future projection are presented in Figure 2.

**Figure 2:**
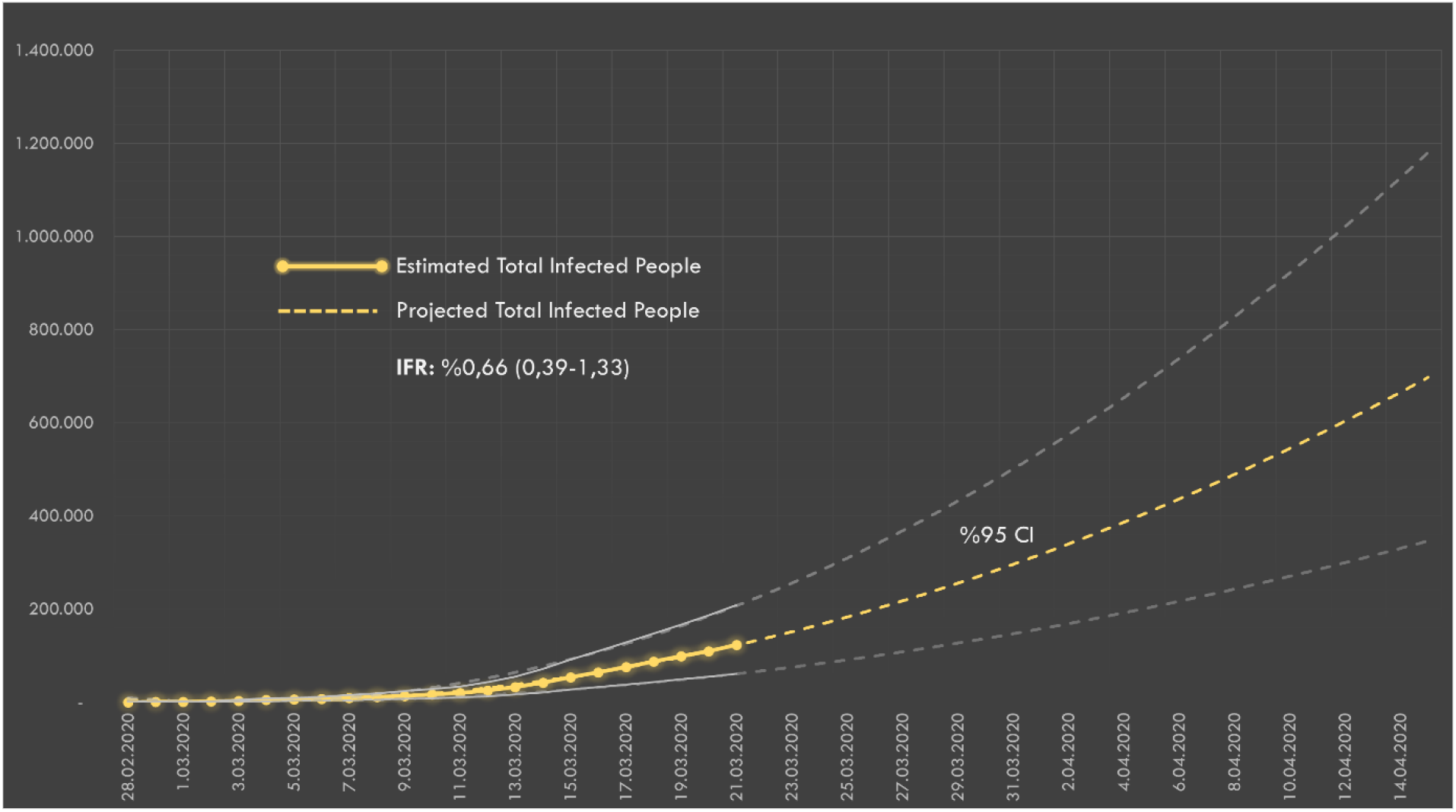
The estimated number of infected people over the number of deaths in Turkey. IFR: Infection Fatality Rate

### 2. Second Dimension

In the case of the free spread of the pandemic without any interventions, the expected age-stratified distribution of the maximum total number of cases, total need for ICU and non-ICU beds and deaths are presented in Figure 3. The maximum total number of hospitalizations was estimated to be 3,418,398, intensive care hospitalizations to be 856,422 and deaths to be 414,203.

**Figure 3:**
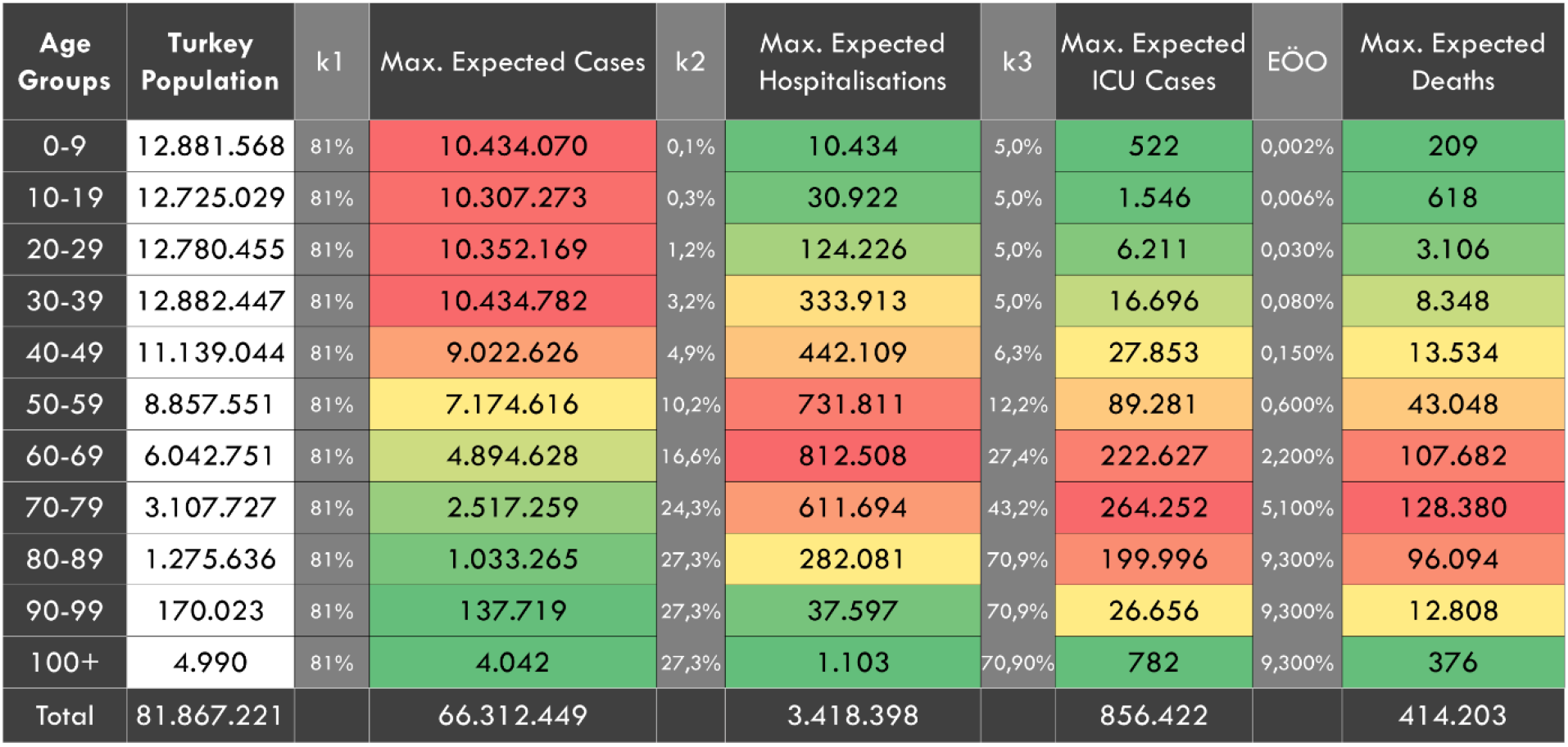
In the case of no interventions, the expected age-stratified distribution of the maximum total case, hospitalization, ICU cases and deaths. k1: Attack rate. k2: age-specific proportions of hospitalization among symptomatic cases. k3: age-specific proportions of ICU need among hospitalized people. IFR: Infection Fatality Rate

### 3. Third Dimension

#### • Scenario 1: No Intervention

The estimations in the second dimension were also simulated in SEIR-based TURKSAS simulator (Table 2). The expected total number of infected people was 72,091,595 and the total number of deaths was 445,956. The attack rate was 88.1% for a pandemic period as the entire society is considered to be the population at risk. The expected mortality ratio was 0·54%.

**Table 2:** Predictions for the first scenario (in the case of no intervention)

|  | Value | Unit |
| --- | --- | --- |
| Expected total cases | 72,091,595 | cases |
| Attack rate | 88.1 | % |
| Expected total deaths | 445,956 | Deaths |
| Mortality | 0.54 | % |
| Daily occupied ICU beds peak | 168,790 | beds |
| Date of peak | June 2020 | date |
| ICU bed capacity exceeded | 4.44 | fold |
| Date ICU beds are 100% full | May 2020 | date |
| Daily occupied non-ICU bed peak | 618,928 | beds |
| Date of peak | June 2020 | date |
| Non-ICU bed capacity exceeded | 3.21 | Fold |
| Date non-ICU beds are 100% full | May 2020 | date |

ICU: Intensive Care Unit

It is predicted that all ICU beds and non-ICU beds will reach 100% occupancy rate in May, while the need for ICU and non-ICU beds would reach its peak in June. At the peak point, the ICU bed capacity would be exceeded by 4·4-fold and the non-ICU bed capacity by 3·21-fold (Figure 4).

**Figure 4:**
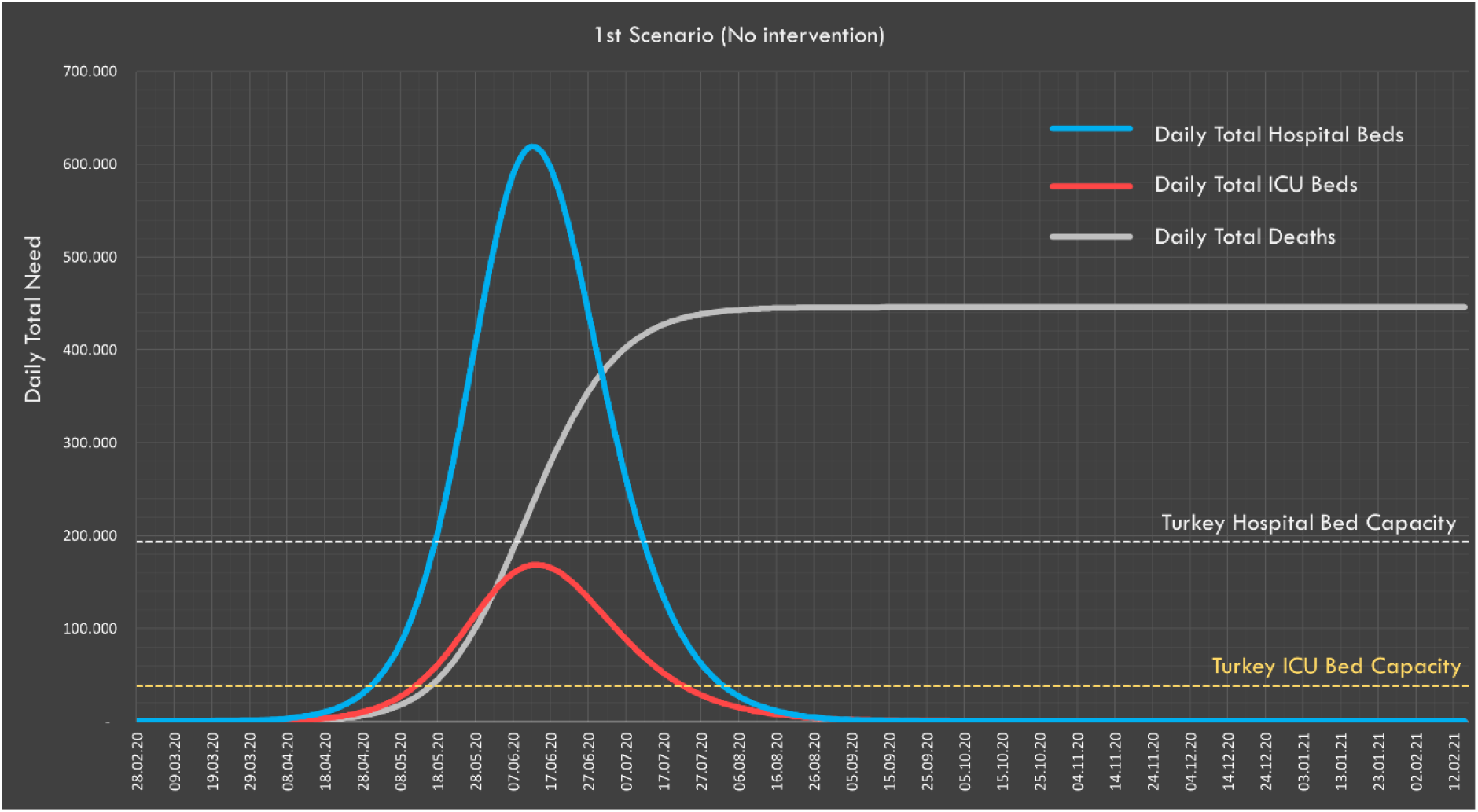
In the worst-case scenario, the need for ICU and non-ICU beds and daily distribution of expected deaths

#### • Scenarios 2 and 3: Social compliance to NPIs (< 100% compliance and 100% compliance)

The effects of the NPIs applied in Turkey on R0 are presented in Figure 5. According to the calculations made by taking into account the compliance rates with the interventions, the value of R0 is estimated to decrease from 3 to 1·38.

**Figure 5:**
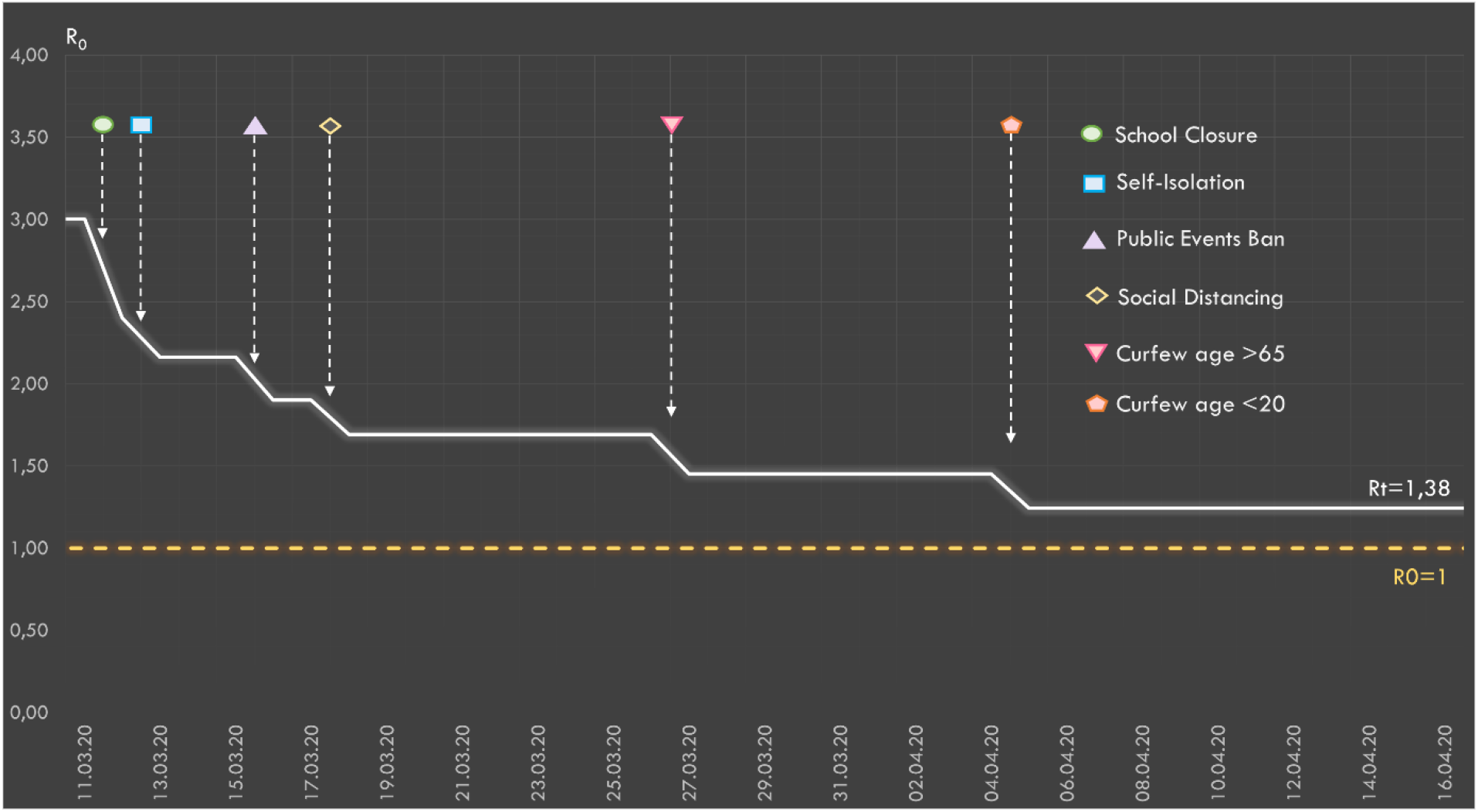
The relative effect of the social interventions applied in Turkey on R0 values

Predictions in first scenario (< 100% compliance) and the second scenario (100% compliance) are presented in Table 3. Compliance with social interventions makes a 94,303 difference in the expected number of deaths. In both scenarios, the predicted peak value of occupied ICU and non-ICU beds remains below Turkey’s healthcare capacity.

**Table 3:**
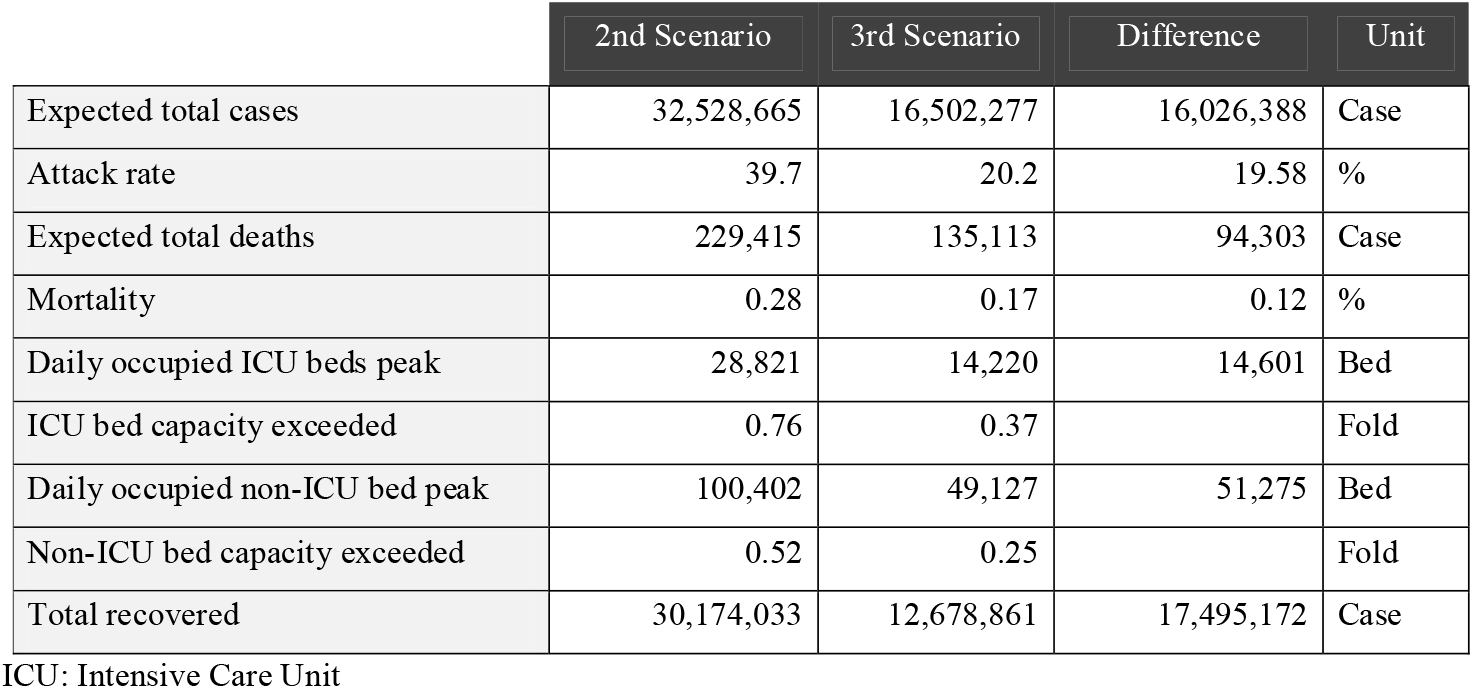
Predictions for the second scenario (< 100% social compliance) and the third scenario (100% social compliance)

For the second and third scenarios, the predicted numbers of total daily deaths and required ICU and non-ICU beds are presented in Figure 6.

**Figure 6:**
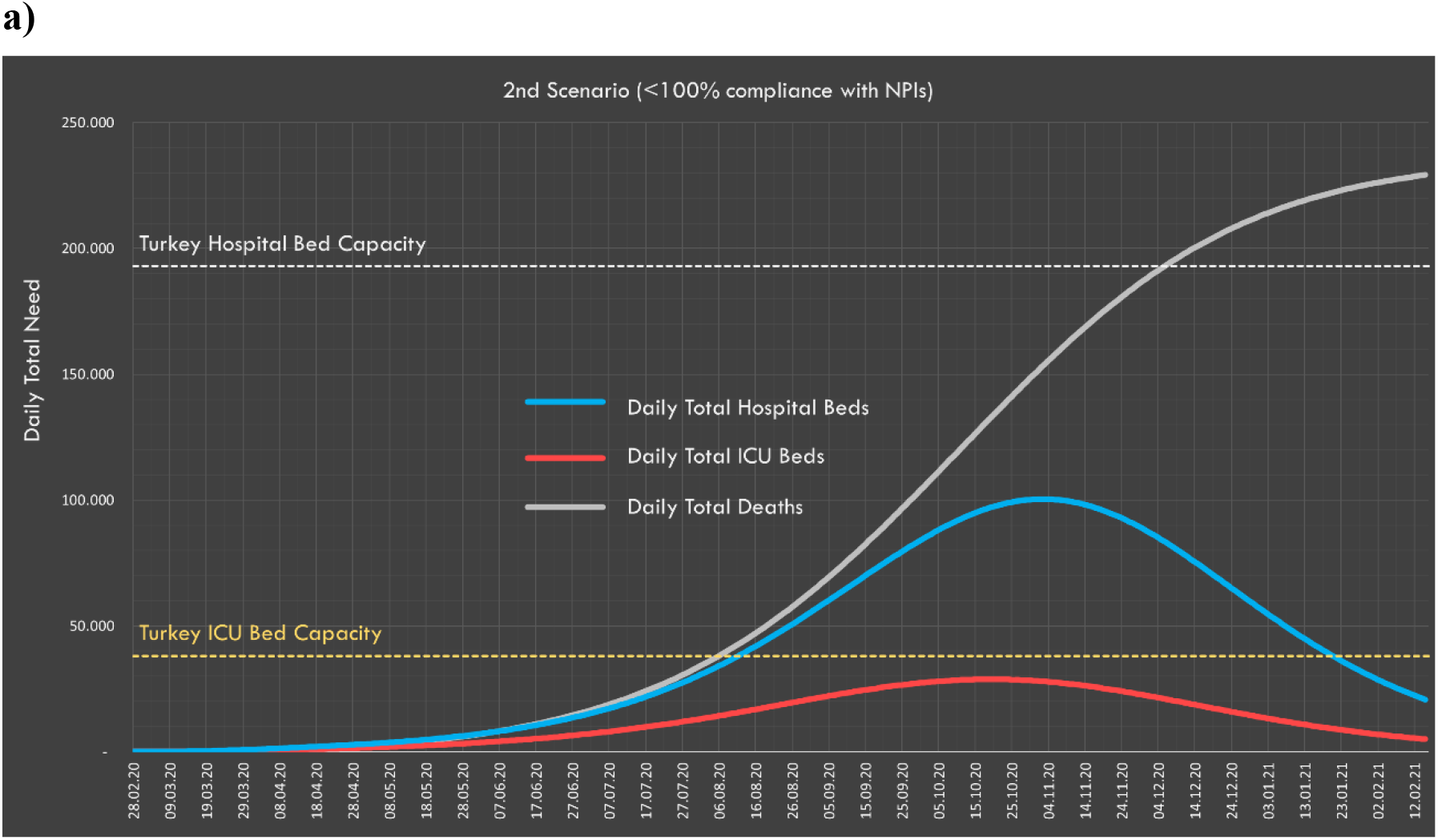
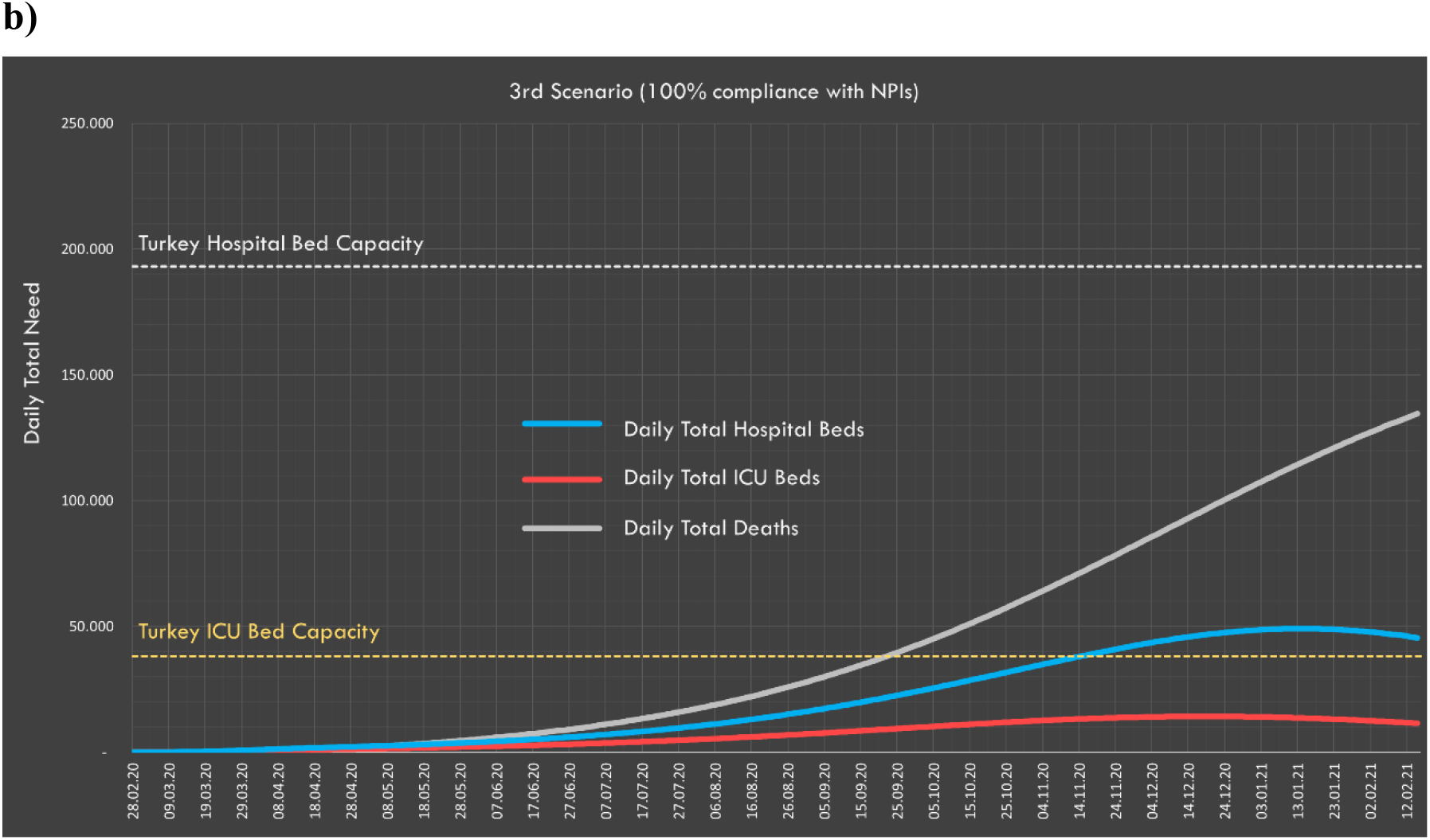
Daily distribution of total ICU and non-ICU beds and expected deaths for the **a)** second and **b)** third scenarios

#### • Scenario 4: General curfew intervention

We predicted that if a curfew were declared for the 21-64 age group, R0 would drop to just below 1 (0·98) and the pandemic would tend towards an end (i.e., non-exponential increase in infection rate). The predicted situation if such a curfew was applied for the 21-64 age group on April 15 is presented in Table 4 and Figure 7. According to these predictions, the expected number of deaths would be 14,230 and the peak daily values for ICU and non-ICU bed demand would be well below the country’s healthcare capacity.

**Table 4:** Predictions for the fourth scenario (general curfew intervention)

|  | Value | Unit |
| --- | --- | --- |
| Expected total cases | 594,924 | Case |
| Attack rate | 0.7 | % |
| Expected total deaths | 14,230 | Deaths |
| Mortality | 0.02 | % |
| Daily occupied ICU beds peak | 1,355 | Beds |
| Date of peak | May 2020 | Date |
| ICU bed capacity exceeded | 0.04 | Fold |
| Daily occupied non-ICU bed peak | 2,146 | Beds |
| Date of peak | May 2020 | Date |

**Figure 7:**
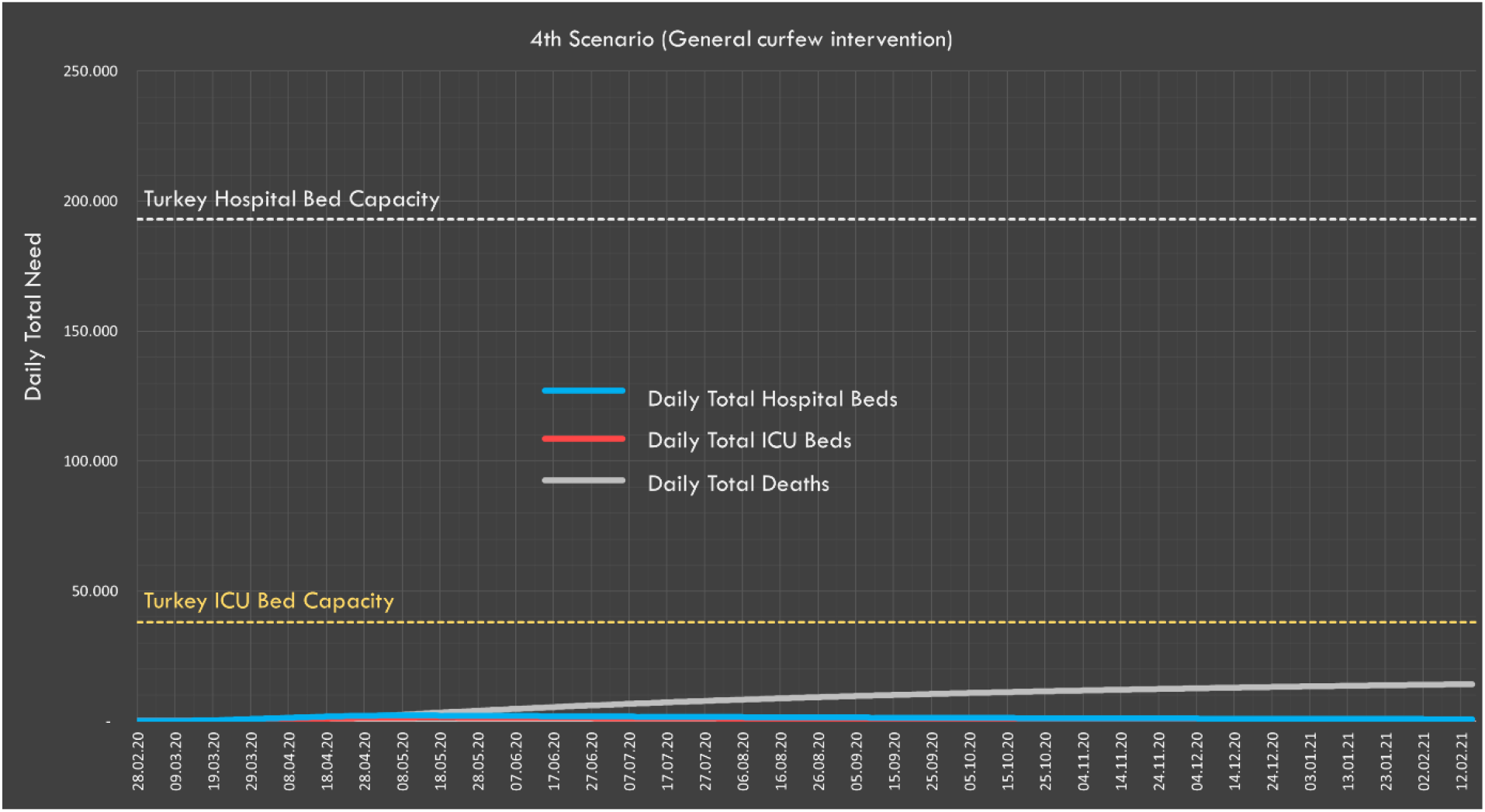
In the fourth scenario, expected daily hospital and ICU bed demand, and distribution of deaths

## Discussion

Estimating and predicting the burden of epidemic diseases on society and the healthcare system in the most accurate way possible is important to ensure the efficient use of the health resources. Although expert opinions are valuable for the predictions relating to the pandemic, it is difficult to find up-to-date evidence to support expert opinions in pandemics that are not frequently experienced. Due to the devastating social effects of epidemics, there is no possibility of experimenting for most interventions, and there are also, of course, ethical limitations involved. For this reason, modelling outbreaks using assumptions supported by the scientific literature and establishing decision support systems based on objective criteria is an important, if not vital, requirement.^26^

### 1. First dimension

The first dimension of the study is to nowcast the actual number of infected people using the IFR. In the estimation of the actual number of cases, the case fatality rate (CFR) and IFR concepts are often confused. The CFR refers to the ratio of the number of deaths in a given time segment to diagnosed cases. However, this rate includes only those who are admitted to hospital and who have been identified, not the proportion of infected people in the community. If perfect conditions were observed and all patients could be followed, how many infected people would die is expressed by the IFR.^13^ For this reason, it is more appropriate to use IFR in the estimation of the final number of deaths and CFR to estimate the number of deaths in a given time period.^14^

We estimated the number of cases in Turkey as 120,000 on 21 March. According to the ICL report, this number was 7 million for Spain as of 28 March 2020; 5·9 million for Italy and 600,000 for Germany.^15^

### 2. Second dimension

In this dimension, the maximum number of infected people was estimated to be 66 million, the number of deaths 414,000 with a consequent mortality rate of 0·54%. According to scientific data for the population of Turkey, this would not be expected to be worse than these numbers.

### 3. Third dimension

In SEIR-based studies, generally, asymptomatic and symptomatic cases have not previously been differentiated according to incubation time, infectivity time and R0 variables. In this study, these two groups were included in the model separately. The proportion of asymptomatic cases can be up to 78% in the studies performed according to the symptoms on the day the PCR sample was taken.^27,28^ However, the WHO stated that 75% of cases that were asymptomatic developed some symptoms at a later stage and the wholly asymptomatic proportion of the population is actually quite low, and therefore not a major determinant of the pandemic ^29^ In the study conducted on the *Diamond Princess*, 17·9% of all cases were stated to be asymptomatic.^19^ In our study, it was accepted that the closest study to the cohort design was the *Diamond Princess* and this same percentage was accordingly used in our calculations. Unlike previous studies, the R compartment was structured with the addition of clinical dynamics in order to evaluate the associated healthcare needs.

In the third dimension of the study, according to this worst-case scenario, a total of 72 million people would be infected in Turkey, and 446,000 people would be estimated to die. According to the ICL report, if there is no intervention, 510,000 deaths would be expected in the UK and 2·2 million in the United States. Also, it is calculated that the ICU bed capacity would be exceeded by 30-fold for the UK.^15^ In our study, the ICU bed capacity in Turkey would be expected to be exceeded by 4·4-fold.

In the second and third scenarios, the expected number of cases and deaths were also calculated according to whether society is partially (second scenario) or fully (third scenario) compliant with the social interventions applied. Predictions show that around 16 million people can be prevented from being infected and 94,000 deaths can be prevented by full compliance with the measures taken. With the measures that Turkey has taken so far, the highest expected need for ICU beds would be under the existing capacity, and indeed ICU bed capacity would not be exceeded were either of these scenarios to be realized. In the fourth scenario, with the implementation of a general curfew that covered all age groups, it was predicted that the total number of cases will be 600,000 and the number of deaths would be less than 15,000.

In our study, we estimate that R0 has decreased to 1·38 as a result of the existing measures in Turkey. This decreases the rate of spread and attack rate of the pandemic. However, in the case of no intervention the attack rate would be 88·1%, while in the case of a general curfew and other NPIs, this value would decrease to 0·7% and overall mortality rates would decline from 0·54% to 0·02%. Complete control of the pandemic is possible by keeping R0 below 1. For this, additional measures would clearly be needed.

In our study, deaths due to exceeding the number of ICU and non-ICU beds were not considered. Also, in case of exceeding intensive care and healthcare capacity, deaths that may result from disruption of healthcare services are not included in the calculations.

Considering that many global and local parameters affect the results, it is quite difficult to draw definitive conclusions or to make clear statements about the natural course of the disease. Mathematical models are important tools in this period where rapid and evidence-based political decisions should be made under the already devastating effects - and potential future effects - of the epidemic. The estimates in this study show that the progressive stages of the pandemic should be carefully projected, and intervention strategies should be evidence-based. The ultimate goal of all NPIs is to maintain the number of cases within the limits that the relevant healthcare system(s) can intervene with until any vaccine or medical treatment method is available, thereby minimizing deaths and disabilities by providing healthcare to as many patients as possible.

Ethical, legal and economic dimensions were ignored in the suggestions presented in this study. The applicability of widespread interventions, which concern not only health but also the economy and social life, should be evaluated through studies in these areas.

## Data Availability

We used external data from:
1) TurkStat data
2) Turkey Health Statistics

http://tuik.gov.tr/PreIstatistikTablo.do?istab_id=1632

https://www.saglik.gov.tr/TR,62400/saglik-istatistikleri-yilligi-2018-yayinlanmistir.html

